# Effect of working from home on the association between job demands and psychological distress

**DOI:** 10.1101/2022.03.10.22272174

**Authors:** Hisashi Eguchi, Akiomi Inoue, Ayako Hino, Mayumi Tsuji, Seiichiro Tateishi, Kazuhiro Ikegami, Tomohisa Nagata, Ryutaro Matsugaki, Yoshihisa Fujino, the CORoNaWork project

**Affiliations:** Department of Mental Health, Institute of Industrial Ecological Sciences, University of Occupational and Environmental Health, Japan, Kitakyushu, Japan; Institutional Research Center, University of Occupational and Environmental Health, Japan, Kitakyushu, Japan; Department of Environmental Health, School of Medicine, University of Occupational and Environmental Health, Japan, Kitakyushu, Japan; Disaster Occupational Health Center, Institute of Industrial Ecological Sciences, University of Occupational and Environmental Health, Kitakyushu, Japan; Department of Work Systems and Health, Institute of Industrial Ecological Sciences, University of Occupational and Environmental Health, Japan, Kitakyushu, Japan; Department of Occupational Health Practice and Management, Institute of Industrial Ecological Sciences, University of Occupational and Environmental Health, Japan, Kitakyushu, Japan; Department of Preventive Medicine and Community Health, School of Medicine, University of Occupational and Environmental Health, Japan, Kitakyushu, Japan; Department of Environmental Epidemiology, Institute of Industrial Ecological Sciences, University of Occupational and Environmental Health, Japan, Kitakyushu, Japan

**Keywords:** COVID-19 pandemic, psychological distress, psychosocial factors, work from home, Japan

## Abstract

**Purpose:** Limited information is available about the association between workplace psychosocial factors and general mental health status among workers during the COVID-19 pandemic. This study examined how working from home affected the association between job demands and psychological distress (PD).

**Method:** A cross-sectional online survey was conducted in December 2020 (N=27,036). The dependent variable (PD) was assessed using the Kessler Psychological Distress Scale. Job demands were assessed using the Job Content Questionnaire. Working from home was determined by participants’ responses to the question: “Do you currently work from home?” We used a two-level regression analysis adjusted for prefecture; each individual-level variable at level 1 was nested into each prefecture at level 2, stratified by working from home or not.

**Results:** Overall, 21.3% of participants worked from home. The interaction between working from home and job demands was significant. Job demands were positively associated with PD. The stratified analysis showed the associations were weaker among employees who worked from home compared with those who did not.

**Conclusion:** The association between job demands and PD may be weakened by working from home.

## Introduction

The ongoing coronavirus disease 2019 (COVID-19) pandemic poses a threat to psychological health. Previous research revealed profound and wide ranging psychosocial impacts at the individual, community, and international levels during past outbreaks of infectious diseases (Mukhtar 2020). The current health crisis has had unprecedented impacts on workplace practices.

High job demands cause depression, which can lead to suicide. Karoshi, or death from overwork, represents a growing public health issue in East Asia (Eguchi et al. 2016). Factors related to COVID-19 infection in the workplace may affect individual coping styles and response to threats (Eguchi et al. 2021). However, workplace-based COVID-19 pandemic countermeasures may weaken the association between job demands and psychological distress.

The COVID-19 pandemic meant that millions of employees across different countries rapidly shifted to working from home. Working from home became crucial for many companies and governments as it allowed people to continue working during the pandemic while reducing the spread of the virus (Şentürk et al. 2021). The positive impacts of working from home, such as reduced commuting time and costs, reduced environmental pollution, and opportunity to support family duties (e.g., picking up children from school) may be desirable for many workers (Kotera and Correa Vione 2020). Employees who worked from home at least 1 day per week demonstrated higher autonomy than those that did not and achieved higher levels of flow (enjoyment, absorption, and intrinsic motivation), which was improved by perceived supervisor and collegial support (Peters et al. 2014). In addition, the association between telecommuting and psychological distress differs depending on telecommuting preference (Otsuka et al. 2021).

The job demands–resources model conceptualizes job resources as “those physical, psychological, social, or organizational aspects of the job that are either/or: 1. functional in achieving work goals, 2. reduce job demands and the associated physiological and psychological costs, and 3. stimulate personal growth, learning, and development” (Demerouti and Bakker 2011, p.2). Working from home may therefore be positioned as a job resource related to job control.

The present study hypothesized that working from home may weaken the association between high job demands and increased psychological distress among general Japanese workers during the third wave of COVID-19 infections. Figure 1 shows the concept model underlying the study hypothesis.

**Figure 1.**
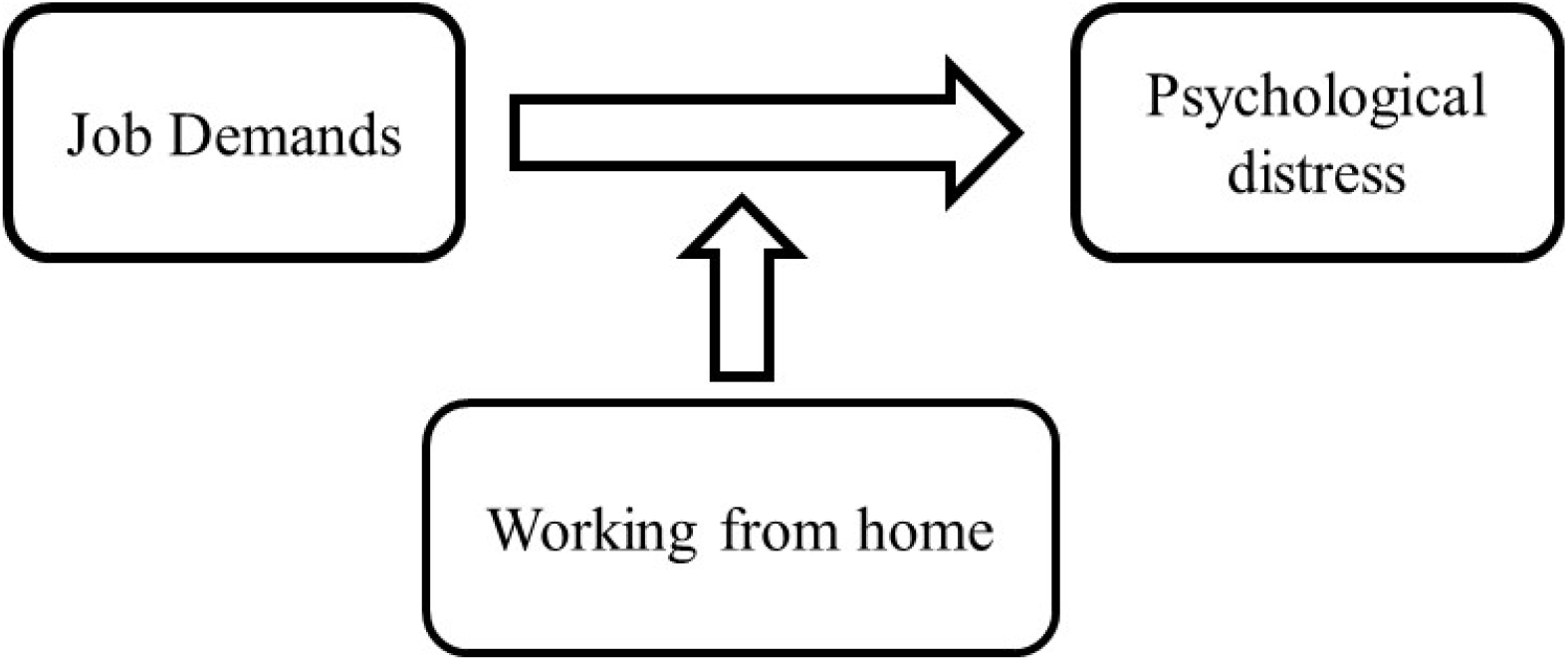
Conceptual model of possible associations between working from home, job demand, and psychological distress

## Materials and Methods

A cross-sectional online survey was conducted in December 2020 among participants who had previously registered with a Japanese web survey company. Invitations to participate were sent to 665,381 registrants via email. Details of the survey protocol have previously been reported (Fujino et al. 2021). A sampling plan was designed to recruit an equal number of respondents from 20 collection units comprising a combination of five regions each, with comparable sex and office/non-office worker status. The target sample size was 1,500 respondents from each collection unit, giving a total of 30,000 respondents. In total, 1,650 respondents (target sample size plus a margin of 10%) were recruited from each collection unit. Of the 33,302 eligible respondents, 215 were excluded because they were deemed to have provided fraudulent responses by Cross Marketing Inc., leaving 33,087 respondents. Participants were selected using a random number generator. The study population comprised individuals interested in participating in a survey. There was a modest financial incentive for survey participation (equivalent to a few US dollars). We excluded 6,051 surveys with invalid responses or response errors, leaving 27,036 surveys for analysis in this study. The exclusion criteria were: extremely short response time (≤6 minutes), extremely low body weight (<30 kg), extremely short height (<140 cm), inconsistent answers to similar questions throughout the survey (e.g., inconsistent responses to questions about marital status and living area), and incorrect answers to a staged question used to identify fraudulent responses (i.e., “Choose the third-largest number from the following five numbers”).

The study aims and protocol were approved by the Ethics Committee of Medical Research, University of Occupational and Environmental Health, Japan (R2-079). Informed consent to participate in this study was obtained from all participants. Participants were informed in advance that their participation was strictly voluntary and all information they provided would remain confidential. Individuals who consented to participate were able to access a designated website (after confirmation of their personal information) where they could complete the survey. Participants had the option to not respond to any part of the questionnaire and could discontinue participation at any time.

### Dependent variable: psychological distress

Psychological distress was assessed using the Kessler Psychological Distress Scale (K6). The K6 was originally developed as a screening instrument for non-specific psychological distress and serious mental illness. Its internal reliability and validity have been documented (Furukawa et al. 2008). The K6 comprises a six-item battery asking how frequently respondents had experienced specific symptoms of psychological distress in the past 30 days. Responses range from 0 (none of the time) to 4 (all of the time), giving a total score of 0–24. The K6 has been translated into Japanese, and the Japanese version has been validated. In this sample, the Cronbach’s α coefficient for the K6 was 0.88.

### Independent variable: Job demands

We used the job demands scale from the Japanese version of the Job Content Questionnaire (JCQ) (Kawakami et al. 1995). The JCQ was developed by Karasek and is based on the job demands– control (or demand–control–support) model. It contains five items that assess job demands, rated on a 4-point scale (1 = strongly disagree to 4 = strongly agree). The total score was calculated according to the JCQ User’s Guide (score range: 12–48) (Karasek et al. 1998). The Japanese version of the JCQ had acceptable reliability and validity (Kawakami et al. 1995). In the present study, the Cronbach’s α coefficient for job demands was 0.68.

### Moderator variable: working from home

Working from home was determined by participants’ responses to the question: “Do you currently work from home?” Response options were “More than 4 days per week,” “More than 2 days per week,” “Less than 1 day per week,” and “Hardly ever.” Responses were subsequently dichotomized using a two-point scale: 0 = yes (“More than 4 days per week,” “More than 2 days per week,” “Less than 1 day per week”); and 1 = no (“Hardly ever”).

### Assessment of covariates

Covariates were measured using a self-administered questionnaire and included demographic and lifestyle characteristics such as sex, age, marital status, educational attainment, occupation, job type, annual family income, and company size. Age was expressed as a continuous variable. Marital status was classified into three categories: married, divorced/widowed, and unmarried. Educational attainment was classified into three categories: junior high school and high school, college and technical school, and university and graduate school. Occupation was classified into 10 categories: staff member; manager; executive; public official/teaching staff/non-profit organization employee; temporary and contract employee; self-employed person; small office/home office worker; agriculture, forestry, and fishery worker; professional (e.g., lawyer, accountant, medical doctor); and others. Job type was classified into three categories: mainly desk work (clerical or computer work), mainly talking to people (e.g., customer service, sales, selling), and mainly labor (e.g., work at construction sites, physical work, nursing care). Participants were asked to indicate their yearly equivalent household income by choosing one of five income bands: (i) 47.4–224.5 million JPY; (ii) 225.0–317.5 million JPY; (iii) 318.1–428.7 million JPY; (iv) 433.0–525.0 million JPY; and (v) 530.3–1050.0 million JPY. Company size was categorized into 10 groups by number of employees: 1 (self-employment), 2–4, 5–9, 10–29, 30–49, 50–99, 100–499, 500–999, 1,000–9,999, and ≥10,000 employees. The cumulative incidence rate of COVID-19 infection 1 week before the survey in the residential prefectures was used as a prefecture-level variable. This information was collected from the websites of public institutions.

### Statistical analyses

Student’s t-tests and chi-square tests were used to examine differences in demographic variables and psychological distress between participants who were working from home and those who were not. We used multilevel regression analyses with two levels adjusted for the prefectural level, whereby each individual-level variable at level 1 was nested into each prefecture at level 2. Examination of the interaction between working from home and job demands showed a significant interaction (p = 0.02). To compare the adjusted coefficients by presence or absence of working from home, multiple regression analyses were used to examine the association between job demands and psychological distress stratified by availability of telecommuting. We conducted multiple regression analysis using a crude model (Model 1) and a model adjusted for sex, age, marital status, educational attainment, occupation, job type, annual household income, and company size (Model 2). All analyses were performed using Stata 15SE (StataCorp, College Station, TX, USA), with statistical significance set at p < 0.05.

## Results

Approximately 20% of participants had the opportunity to work from home. Employees who worked from home were older and had lower psychological distress than those who did not work from home. Men, self-employed people, those with a higher household income, those whose job mainly involved desk work, and employees in large companies were more likely to work from home (Table 1).

**Table 1.**
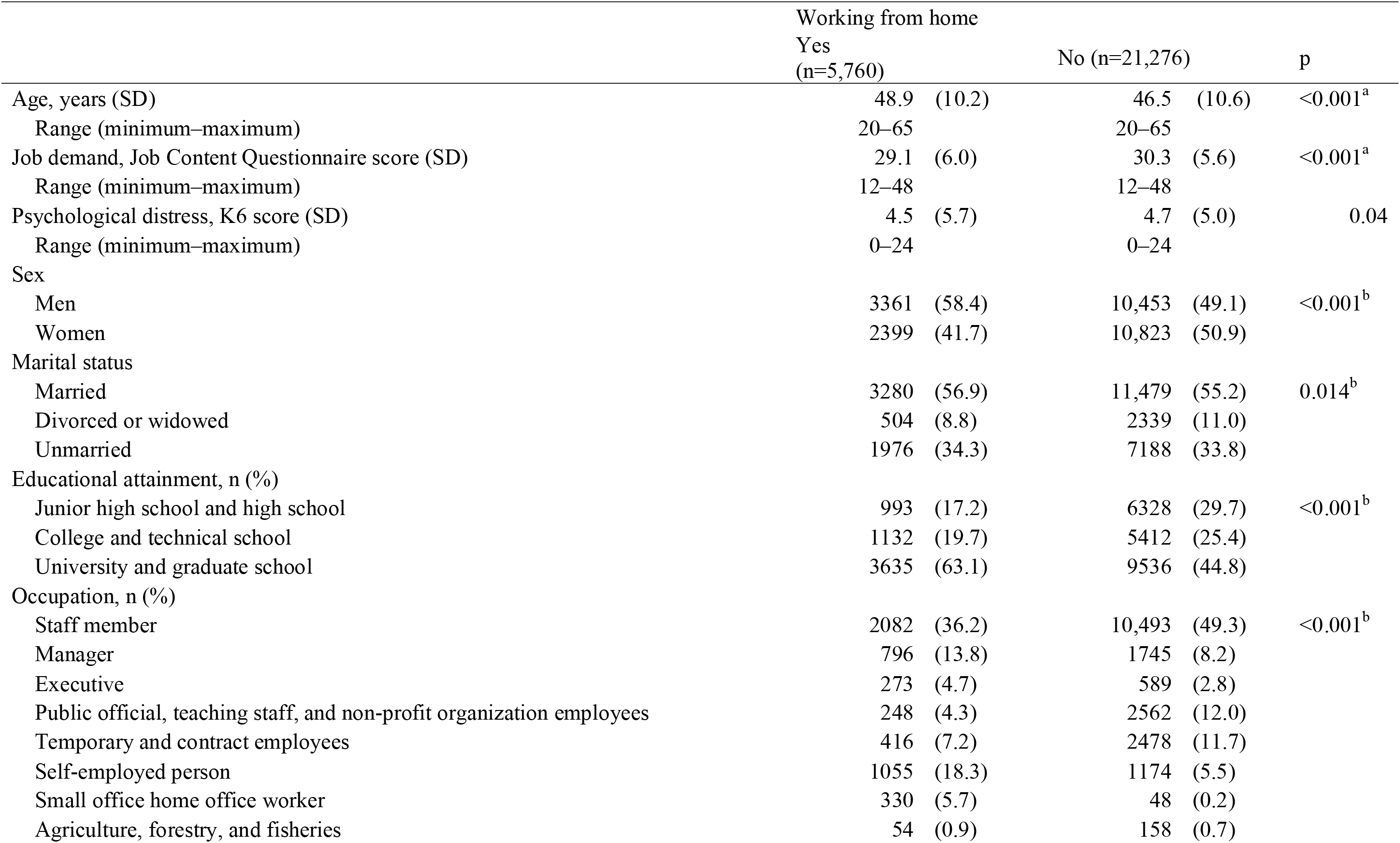
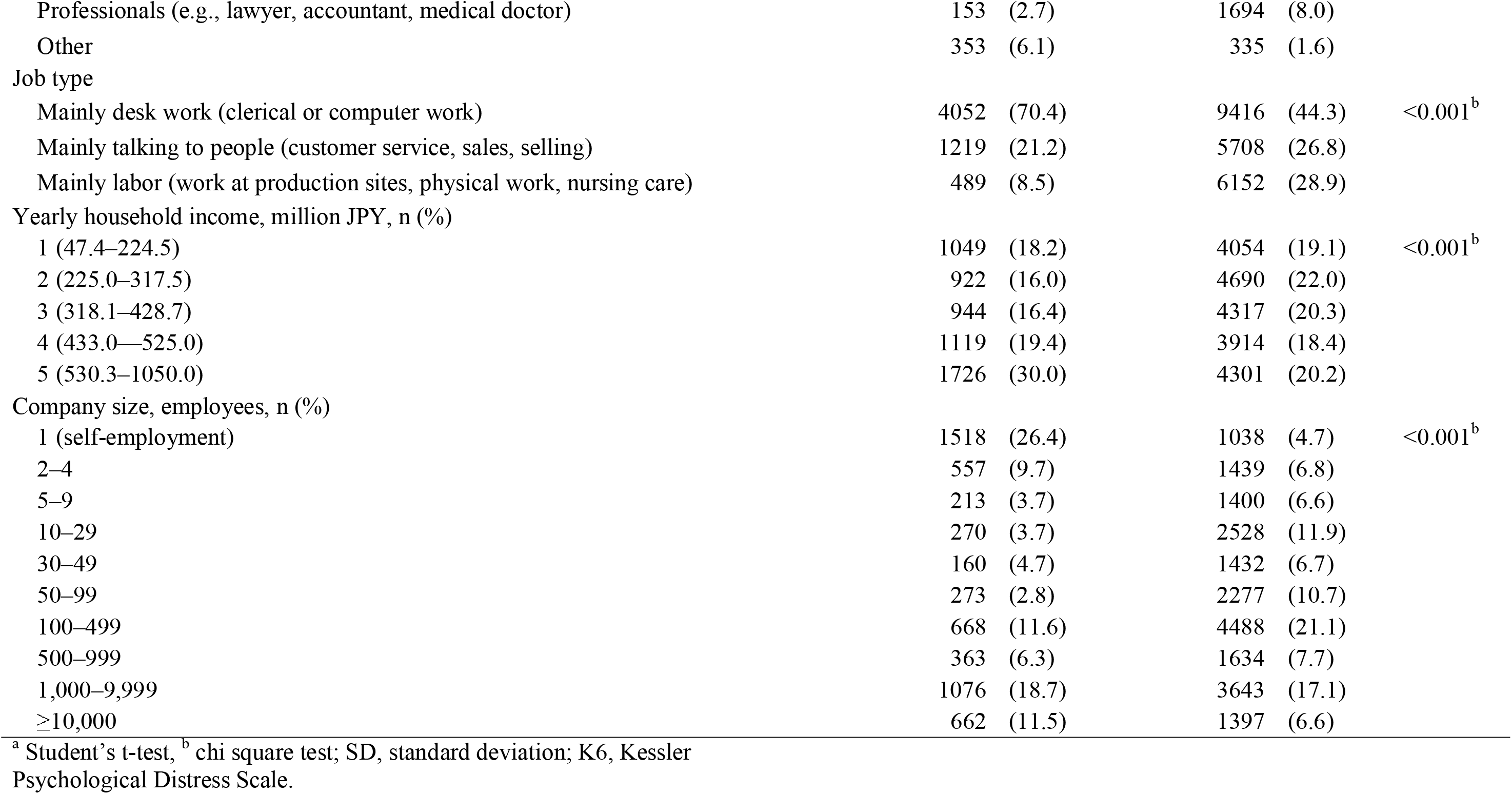
Participants’ characteristics (N=27,036)

|  | Working from home |  |  |  | p |
| --- | --- | --- | --- | --- | --- |
|  | Yes<br>(n=5,760) |  | No (n=21,276) |  |  |
| Age, years (SD) | 48.9 (10.2) |  | 46.5 (10.6) |  | <0.001<sup>a</sup> |
| Range (minimum–maximum) | 20–65 |  | 20–65 |  |  |
| Job demand, Job Content Questionnaire score (SD) | 29.1 (6.0) |  | 30.3 (5.6) |  | <0.001<sup>a</sup> |
| Range (minimum–maximum) | 12–48 |  | 12–48 |  |  |
| Psychological distress, K6 score (SD) | 4.5 (5.7) |  | 4.7 (5.0) |  | 0.04 |
| Range (minimum–maximum) | 0–24 |  | 0–24 |  |  |
| Sex |  |  |  |  |  |
| Men | 3361 (58.4) |  | 10,453 (49.1) |  | <0.001<sup>b</sup> |
| Women | 2399 (41.7) |  | 10,823 (50.9) |  |  |
| Marital status |  |  |  |  |  |
| Married | 3280 (56.9) |  | 11,479 (55.2) |  | 0.014<sup>b</sup> |
| Divorced or widowed | 504 (8.8) |  | 2339 (11.0) |  |  |
| Unmarried | 1976 (34.3) |  | 7188 (33.8) |  |  |
| Educational attainment, n (%) |  |  |  |  |  |
| Junior high school and high school | 993 (17.2) |  | 6328 (29.7) |  | <0.001<sup>b</sup> |
| College and technical school | 1132 (19.7) |  | 5412 (25.4) |  |  |
| University and graduate school | 3635 (63.1) |  | 9536 (44.8) |  |  |
| Occupation, n (%) |  |  |  |  |  |
| Staff member | 2082 (36.2) |  | 10,493 (49.3) |  | <0.001<sup>b</sup> |
| Manager | 796 (13.8) |  | 1745 (8.2) |  |  |
| Executive | 273 (4.7) |  | 589 (2.8) |  |  |
| Public official, teaching staff, and non-profit organization employees | 248 (4.3) |  | 2562 (12.0) |  |  |
| Temporary and contract employees | 416 (7.2) |  | 2478 (11.7) |  |  |
| Self-employed person | 1055 (18.3) |  | 1174 (5.5) |  |  |
| Small office home office worker | 330 (5.7) |  | 48 (0.2) |  |  |
| Agriculture, forestry, and fisheries | 54 (0.9) |  | 158 (0.7) |  |  |

|  |  |  |  |
| --- | --- | --- | --- |
| Professionals (e.g., lawyer, accountant, medical doctor) | 153 (2.7) | 1694 (8.0) |  |
| Other | 353 (6.1) | 335 (1.6) |  |
| Job type |  |  |  |
| Mainly desk work (clerical or computer work) | 4052 (70.4) | 9416 (44.3) | <0.001<sup>b</sup> |
| Mainly talking to people (customer service, sales, selling) | 1219 (21.2) | 5708 (26.8) |  |
| Mainly labor (work at production sites, physical work, nursing care) | 489 (8.5) | 6152 (28.9) |  |
| Yearly household income, million JPY, n (%) |  |  |  |
| 1 (47.4–224.5) | 1049 (18.2) | 4054 (19.1) | <0.001<sup>b</sup> |
| 2 (225.0–317.5) | 922 (16.0) | 4690 (22.0) |  |
| 3 (318.1–428.7) | 944 (16.4) | 4317 (20.3) |  |
| 4 (433.0–525.0) | 1119 (19.4) | 3914 (18.4) |  |
| 5 (530.3–1050.0) | 1726 (30.0) | 4301 (20.2) |  |
| Company size, employees, n (%) |  |  |  |
| 1 (self-employment) | 1518 (26.4) | 1038 (4.7) | <0.001<sup>b</sup> |
| 2–4 | 557 (9.7) | 1439 (6.8) |  |
| 5–9 | 213 (3.7) | 1400 (6.6) |  |
| 10–29 | 270 (3.7) | 2528 (11.9) |  |
| 30–49 | 160 (4.7) | 1432 (6.7) |  |
| 50–99 | 273 (2.8) | 2277 (10.7) |  |
| 100–499 | 668 (11.6) | 4488 (21.1) |  |
| 500–999 | 363 (6.3) | 1634 (7.7) |  |
| 1,000–9,999 | 1076 (18.7) | 3643 (17.1) |  |
| ≥10,000 | 662 (11.5) | 1397 (6.6) |  |

<sup>a</sup> Student's t-test, <sup>b</sup> chi square test; SD, standard deviation; K6, Kessler Psychological Distress Scale.

We demonstrated a significant association between job demands and psychological distress (p < 0.05). The stratified analysis (post hoc simple slope analysis) showed that the effect of job demands on psychological distress was weaker among employees who worked from home (coefficient = 0.15) than among those who did not (coefficient = 0.18) (Table 2). Company size was positively associated with psychological distress among those who worked from home, and negatively associated with psychological distress among those who did not work from home.

**Table 2.**
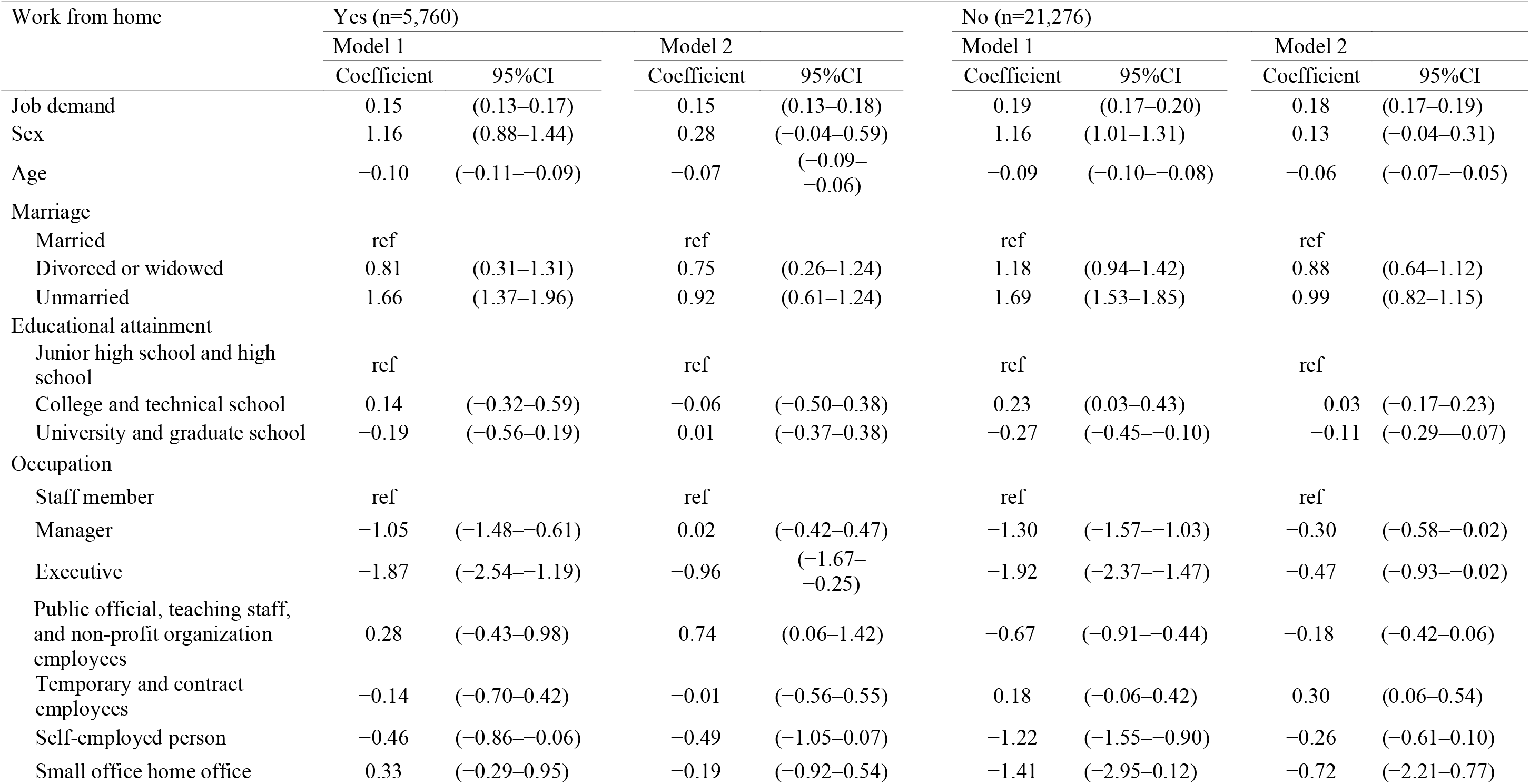
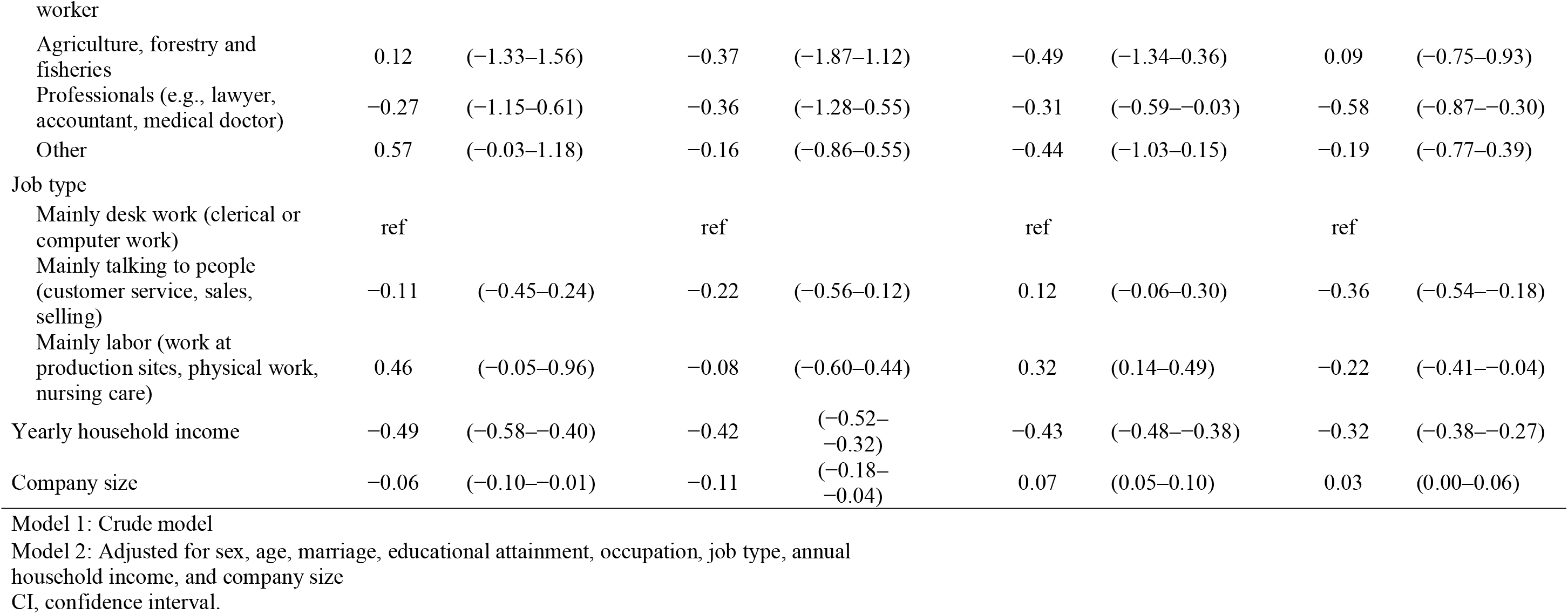
Associations between job demand and psychological distress by working from home (N=27,036)

## Discussion

We conducted a large online survey on December 22, 2020, just before the government declared a second state of emergency in the greater Tokyo area (January 7, 2021). Men, self-employed people, those with a higher household income, those whose job mainly involved desk work, and employees in large companies were more likely to work from home. The association between job demands and psychological distress was stronger among employees who did not work from home than among those worked from home.

Working from home may weaken the association between job demands and psychological distress. Working from home has been associated with reduced commuting time and costs, reduced environmental pollution, and the opportunity to support family duties (e.g., picking up children from school), which may be desirable for many workers (Kotera and Correa Vione 2020). Having a sense of control over worktime can also help employees manage their work-life balance (Beckers et al. 2012). Irrespective of the COVID-19 pandemic, employers may proactively provide opportunities to work from home to prevent psychological distress among employees. In addition, working from home may reduce the fear of infection at or on the way to work (Nguyen 2021; Sato et al. 2021). Our previous study showed that the association between job demands and psychological distress may be strengthened by anxiety about COVID-19 infection in the workplace (Eguchi et al. 2020). Working from home may affect the association between job demands and psychological distress through reducing anxiety about COVID-19 infection.

The effect of company size on psychological distress differed between those who worked from home and those that did not. The association between company size and psychological distress in previous studies was inconsistent (Inoue et al. 2010; Kanamori et al. 2020). This discrepancy may be attributable to the use of different indicators (company size vs. worksite size) or different survey years reflecting different economic situations. Working from home decreased employees’ communication with their supervisors and colleagues (Amano et al. 2021). Therefore, working from home may affect the association between company size and psychological distress via communication changes.

This study had some limitations. First, our study population required Internet access to complete the survey and therefore might have comprised participants that were more aware of COVID-19 infection through access to online information. People should be aware of the psychological risk of too much media exposure and control their access in health crises such as the COVID-19 outbreak (Sasaki et al. 2020). Our results are not completely generalizable to individuals without Internet access or to people in other countries and settings. Second, we had no information about participants’ personality traits. In addition, we had no information about the number of confirmed cases of COVID-19 in the workplace. Further studies are needed to evaluate whether other confounding factors provide possible mechanisms for the observed attenuation in the associations between job demands, working from home, and psychological distress. Third, this study used a cross-sectional design, and no causal associations could be determined. A further study using an interventional or prospective design is needed to clarify potential causal associations between job demands, working from home, and psychological distress in the Japanese working population. Finally, we should consider the possible effects of common method bias when interpreting the results.

## Conclusion

The association between job demands and psychological distress may be weakened by working from home.

## Data Availability

All data produced in the present study are available upon reasonable request to the authors.

## Notes

Conflicts of interest All authors declare that they have no competing interests.

### Competing Interest Statement

The authors have declared no competing interest.

### Funding Statement

This study was funded by: a research grant from the University of Occupational and Environmental Health, Japan, a general incorporated foundation (Anshin Zaidan) for the development of educational materials on mental health measures for managers at small-sized enterprises, Health, Labour and Welfare Sciences Research Grants: Comprehensive Research for Women's Healthcare (H30-josei-ippan-002), Research for the establishment of an occupational health system in times of disaster (H30-roudou-ippan-007) and Work-related Diseases Clinical Research Grant 2021 (200401-01), consigned research foundation (the Collabo-health Study Group), scholarship donations from Chugai Pharmaceutical Co., Ltd., and the Japan Society for the Promotion of Science (JSPS KAKENHI: Grant Number JP20K10477).

### Author Declarations

The Ethics Committee of Medical Research, University of Occupational and Environmental Health, Japan gave ethical approval for this work (R2-079).

